# Suppression of COVID-19 outbreak in the municipality of Vo’, Italy

**DOI:** 10.1101/2020.04.17.20053157

**Authors:** Enrico Lavezzo, Elisa Franchin, Constanze Ciavarella, Gina Cuomo-Dannenburg, Luisa Barzon, Claudia Del Vecchio, Lucia Rossi, Riccardo Manganelli, Arianna Loregian, Nicolò Navarin, Davide Abate, Manuela Sciro, Stefano Merigliano, Ettore Decanale, Maria Cristina Vanuzzo, Francesca Saluzzo, Francesco Onelia, Monia Pacenti, Saverio Parisi, Giovanni Carretta, Daniele Donato, Luciano Flor, Silvia Cocchio, Giulia Masi, Alessandro Sperduti, Lorenzo Cattarino, Renato Salvador, Katy A. M. Gaythorpe, Imperial College London COVID-19 Response Team, Alessandra R. Brazzale, Stefano Toppo, Marta Trevisan, Vincenzo Baldo, Christl A. Donnelly, Neil M. Ferguson, Ilaria Dorigatti, Andrea Crisanti

## Abstract

On the 21^st^ of February 2020 a resident of the municipality of Vo’, a small town near Padua, died of pneumonia due to SARS-CoV-2 infection^1^. This was the first COVID-19 death detected in Italy since the emergence of SARS-CoV-2 in the Chinese city of Wuhan, Hubei province^2^. In response, the regional authorities imposed the lockdown of the whole municipality for 14 days^3^. We collected information on the demography, clinical presentation, hospitalization, contact network and presence of SARS-CoV-2 infection in nasopharyngeal swabs for 85.9% and 71.5% of the population of Vo’ at two consecutive time points. On the first survey, which was conducted around the time the town lockdown started, we found a prevalence of infection of 2.6% (95% confidence interval (CI) 2.1-3.3%). On the second survey, which was conducted at the end of the lockdown, we found a prevalence of 1.2% (95% CI 0.8-1.8%). Notably, 43.2% (95% CI 32.2-54.7%) of the confirmed SARS-CoV-2 infections detected across the two surveys were asymptomatic. The mean serial interval was 6.9 days (95% CI 2.6-13.4). We found no statistically significant difference in the viral load (as measured by genome equivalents inferred from cycle threshold data) of symptomatic versus asymptomatic infections (p-values 0.6 and 0.2 for E and RdRp genes, respectively, Exact Wilcoxon-Mann-Whitney test). Contact tracing of the newly infected cases and transmission chain reconstruction revealed that most new infections in the second survey were infected in the community before the lockdown or from asymptomatic infections living in the same household. This study sheds new light on the frequency of asymptomatic SARS-CoV-2 infection and their infectivity (as measured by the viral load) and provides new insights into its transmission dynamics, the duration of viral load detectability and the efficacy of the implemented control measures.

## Introduction

As of 2^nd^ April 2020, 857,641 confirmed cases and 42,006 deaths of a Novel Coronavirus Disease (COVID-19) have been reported worldwide, affecting 201 countries^2^. In Italy, COVID-19 has caused over 7,500 confirmed deaths, the largest number of deaths in any country. The causative agent (SARS-CoV-2), a close relative of SARS-CoV^4^, was introduced into the human population of Wuhan City, Hubei province (China) around the beginning of December 2019^5,6^. In Hubei province and in the rest of mainland China, the epidemic is being successfully contained with strategies based on the isolation of cases and their contacts, along with drastic social distancing measures that include the quarantine of whole cities and regions, the closure of schools and workplaces and the cancellations of mass gatherings. Recent reports suggest that the drastic interventions implemented in mainland China had a tremendous effect on the control of the epidemic^7,8^. However, the long-term effectiveness of these interventions remains unclear^9^. Sustained transmission is currently ongoing in several countries outside mainland China, with European countries (in particular Italy and Spain), the United States and Iran being most affected^2^. In Europe, unprecedented interventions have been implemented in order to suppress the transmission of SARS-CoV-2. Despite the mounting pressure on healthcare demand and the growing death toll, new analyses suggest that the interventions currently in place will soon start to control the epidemic, bringing hope to all affected countries, including Italy^10^. Several uncertainties currently remain regarding the transmission dynamics of the virus, particularly on the contribution of asymptomatic versus symptomatic infections to the onward transmission of the virus^11^. Effective long-term control of transmission in Europe and worldwide, depends on an improved understanding of the mechanisms of SARS-CoV-2 transmission. This is particularly important given that, in the absence of a vaccine or effective treatment, alternative public health interventions are being evaluated to allow the population to maintain essential societal and economic activities while at the same time controlling the spread of SARS-CoV-2, limiting mortality and maintaining healthcare demand within capacity.

In Italy, the first death linked to COVID-19 occurred on 21^st^ February 2020 in the municipality of Vo’, in the Veneto region^12^. On that occasion, the national and regional authorities enforced the closure of all public services and commercial activities and imposed a ban on population movement from 23^rd^ February 2020 to 8^th^ March 2020. During this period, we tested the entire population twice for the presence of virus in nasopharyngeal swabs. Thus, Vo’ represented a unique opportunity for understanding the epidemiology of SARS-CoV-2 and its transmission dynamics in unprecedented detail. The experience of Vo’ represents an independent proof-of-concept that despite the silent and widespread transmission of SARS-CoV-2, transmission can be suppressed. In this study we present the results of two surveys of the resident population of Vo’, conducted less than two weeks apart, to investigate population exposure to SARS-CoV-2 before and after the lockdown.

We also present an analysis of the data collected within the surveys concerning population demography, prevalence of infection, frequency of symptomatic vs asymptomatic infection and viral load in symptomatic vs asymptomatic infections. We assessed the risk of SARS-CoV-2 infection associated with comorbidity and therapies for underlying conditions, characterised chains of transmission, studied the transmission dynamics of SARS-CoV-2 and assessed the impact of the social distancing measures implemented. Our analyses show that viral transmission could be effectively and rapidly suppressed by combining the early isolation of infected people with community lockdown. This experience represents a model for settings with similar epidemiological and demographic conditions.

## Results

### Study setting

During the two surveys we collected nasopharyngeal swabs from 2,812 and 2,343 subjects, corresponding to 85.9% and 71.5% of the eligible study population (**Figure 1**). All age groups were homogeneously sampled with age-specific percentages ranging from 70.8% to 91.6% in the first survey and 52.4% to 81.6% in the second survey (**Table S1**). Statistical analysis showed that no significant bias was introduced in the composition of the different age groups when comparing the two surveys (Fisher exact test, p-value = 0.20) (**Figure S1**).

**Figure 1:**
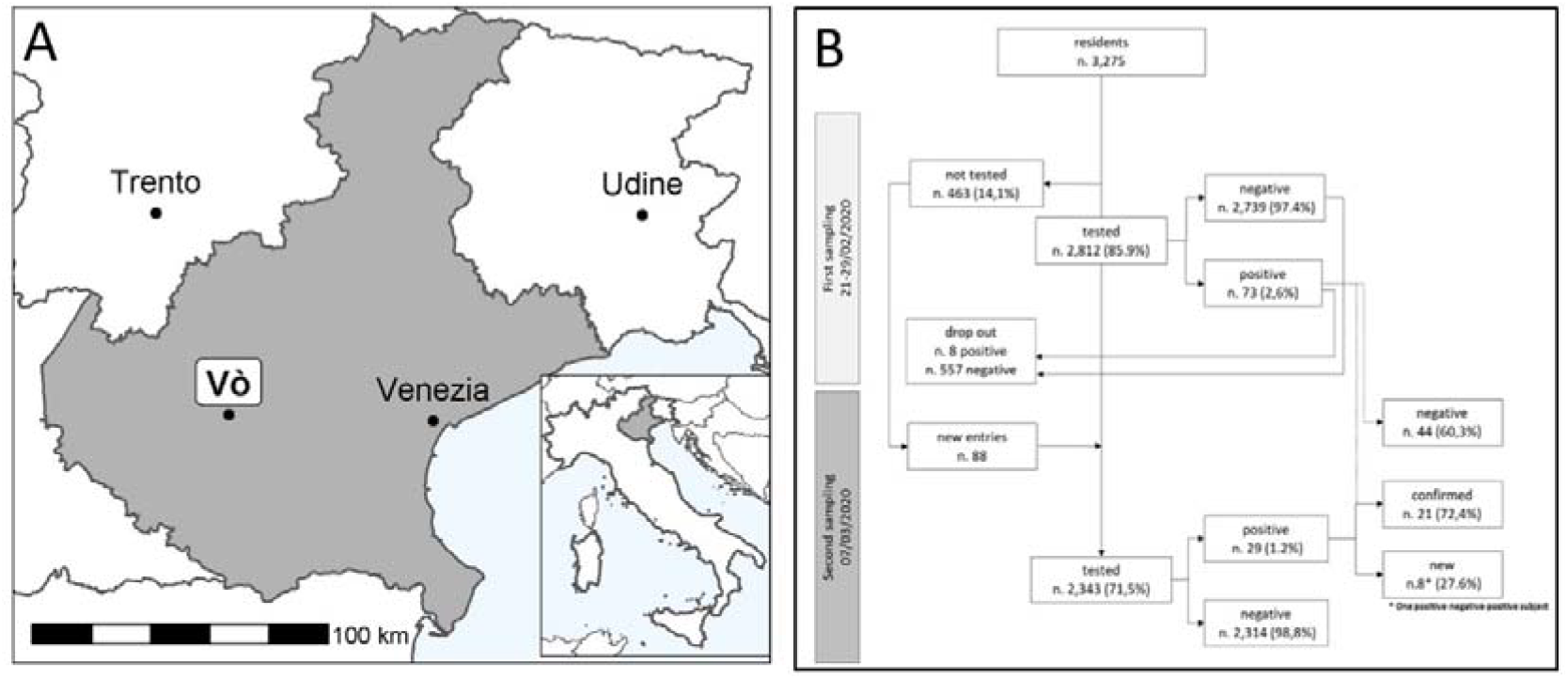
Description of the geography and study setting. (A) Map showing the location of Vo’ and of the Veneto region (grey area) within Italy. (B) Flow chart summarising the key statistics on the two sequential nasopharyngeal swab surveys conducted in Vo’ to assess the transmission of SARS-CoV-2 before and after the implementation of interventions.

### Analysis of infection prevalence

A total of 73 out of the 2,812 subjects tested at the first time-point were positive, which gives a prevalence of 2.6% (95% CI 2.1-3.3%) (**Table 1**). The second survey identified 29 total positive cases (prevalence 1.2%; 95% CI 0.8-1.8%), 8 of which were new cases (0.3%; 95% CI 0.15-0.7%) (**Figure 2**). One of the 8 new infections detected in the second survey is a hospitalized subject who tested positive, then negative, then positive again. An alternative interpretation would be that the second test was a false negative. The frequency of the symptoms in the SARS-CoV-2 positive individuals was systematically recorded, with fever and cough being the most common (**Figure S2**). Notably, a total of 30 out of the 73 individuals (41.1%; 95% CI 29.7-53.2%) who tested positive at the first survey were asymptomatic (i.e. they did not report fever, cough or any other symptoms, according to the definition used in this analysis). A similar proportion of asymptomatic infection was also recorded at the second survey (13 out of 29, 44.8%; 95% CI 26.5-64.3%); in the 8 new cases, 5 were asymptomatic (**Table 2, Figure S3**). No infections were detected in either survey in 234 tested children ranging from 0 to 10 years, despite some of them living in the same household as infected people (**Table S3**). Up to the age of 50 years, the prevalence of infection oscillated between a central estimate of 1.2% to 1.7% (**Figure S4**). Older individuals showed a three-fold increase in the prevalence of infection (**Table 2, Figure S4**). Of the 81 SARS-CoV-2 positive patients across the two surveys, 14 required hospitalization (17.2%). Their age distribution was as follows: 1 (7.1%) in the 41-50 age group, 2 (14.3%) in the 51-60 age group, 4 (28.6%) in the 61-70 age group, 5 (35.7%) in the 71-80 age group and 2 (14.3%) in the 81-90 age group.

**Table 1.**
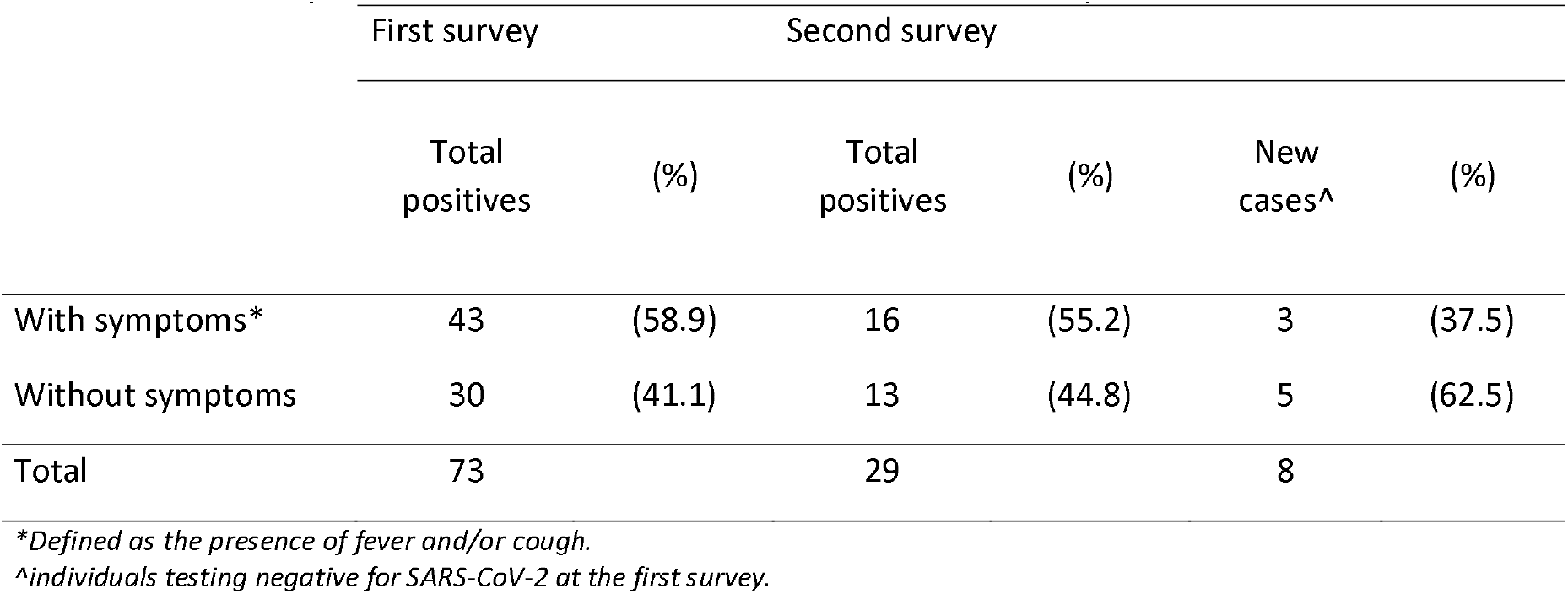
Individuals positive for SARS-CoV-2 at the first and second survey.

**Table 2.** Individuals positive for SARS-CoV-2 at the first and second survey stratified by sex and age groups.

|  |  | First survey |  |  | Second survey |  |  |  |  |
| --- | --- | --- | --- | --- | --- | --- | --- | --- | --- |
|  |  | n | Total | (%) | n | Total | (%) | New cases | (%) |
| Gender |  |  |  |  |  |  |  |  |  |
|  | Males | 1408 | 43 | (3.1) | 1165 | 20 | (1.7) | 5 | (0.4) |
|  | Females | 1404 | 30 | (2.1) | 1178 | 9 | (0.8) | 3 | (0.3) |
|  | p-value |  |  | 0.15 |  |  | 0.041 |  |  |
| Age group |  |  |  |  |  |  |  |  |  |
|  | 00-10 | 217 | 0 | (0.0) | 157 | 0 | (0.0) |  | (0.0) |
|  | 11-20 | 250 | 3 | (1.2) | 210 | 2 | (1.0) | 1 | (0.5) |
|  | 21-30 | 240 | 4 | (1.7) | 191 | 2 | (1.0) |  | (0.0) |
|  | 31-40 | 286 | 7 | (2.4) | 241 | 2 | (0.8) |  | (0.0) |
|  | 41-50 | 439 | 5 | (1.1) | 366 | 2 | (0.5) | 1 | (0.3) |
|  | 51-60 | 496 | 16 | (3.2) | 439 | 7 | (1.6) | 2 | (0.5) |
|  | 61-70 | 384 | 15 | (3.9) | 349 | 6 | (1.7) | 2 | (0.6) |
|  | 71-80 | 318 | 19 | (6.0) | 262 | 6 | (2.3) | 2 | (0.8) |
|  | 81+ | 182 | 4 | (2.2) | 128 | 2 | (1.6) |  | (0.0) |
|  | p-value |  |  | < 0.001* |  |  | 0.48 |  |  |
| Total |  | 2,812 | 73 | (2.6) | 2,343 | 29 | (1.2) | 8 | (0.3) |

P-values were computed using Fisher's exact test (for gender) and the likelihood ratio test (for age-group). \*Linear trend.

**Figure 2:**
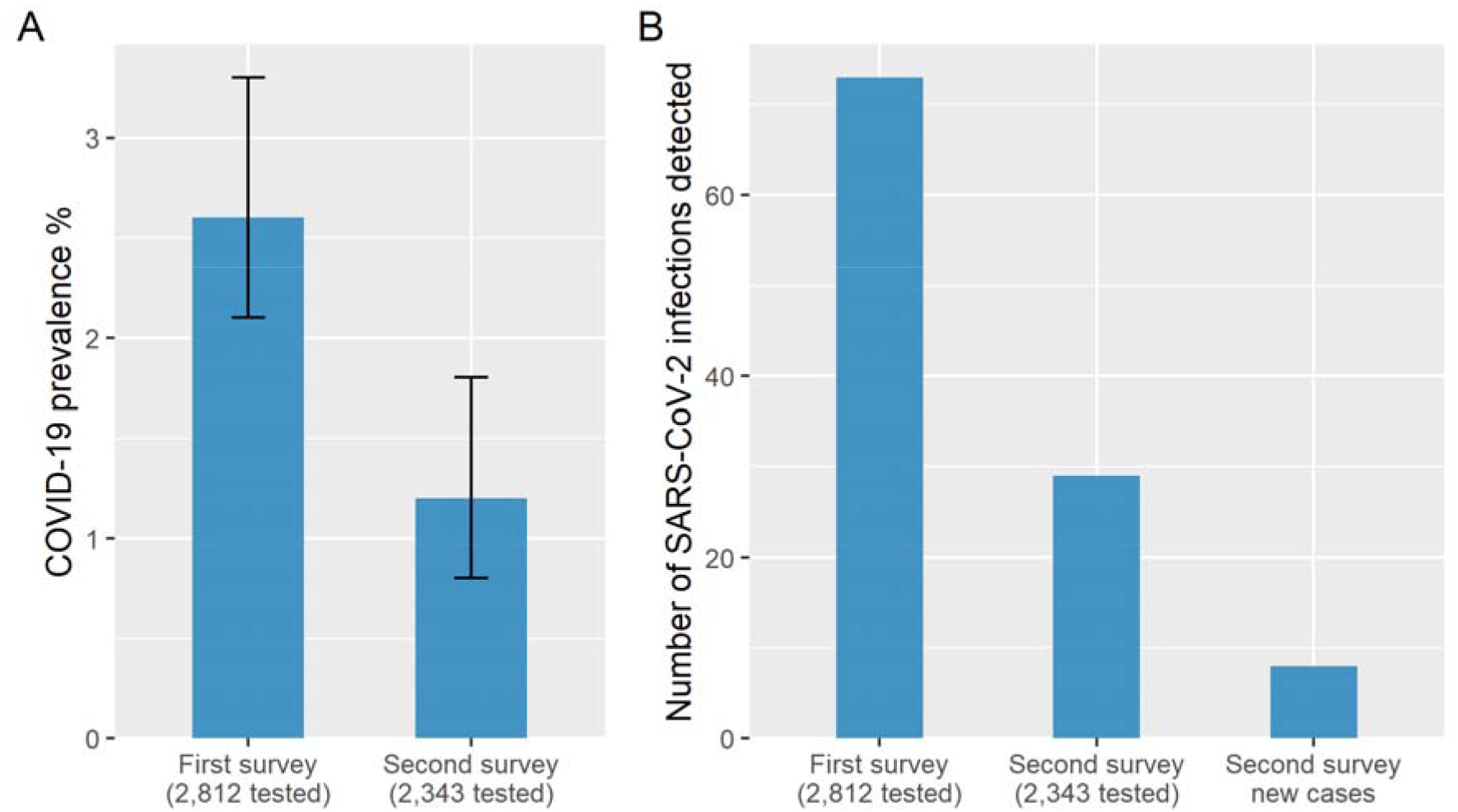
**(A)** Prevalence of SARS-CoV-2 infection at the first and second survey. (B) Number of SARS-CoV-2 infections detected among the sampled population of the residents of Vo’ in the first and in the second survey.

A substantial fraction of infected individuals (67.7%; 95% CI 54.9%-78.8%, symptomatic and asymptomatic combined over all ages) cleared the infection between the first and second surveys (**Table S2**). The time to viral clearance (time from the earliest positive sample for the subjects with more than one sample in the first survey and a negative sample in the second survey) ranged from 8 to 13 days and was on average 9.3 days, with standard deviation 2.0 days. The minimal duration of the positivity window (time from the earliest positive sample for the subjects with more than one sample in the first survey and a positive sample in the second survey) ranged from 3 to 13 days and was on average 9.1 days, with standard deviation 2.3 days. In particular, 68.3% (95% CI 51.9-81.9%) of symptomatic and 66.7% (95% CI 44.7-84.4%) of asymptomatic SARS-CoV-2 infections cleared the virus during the study period (i.e. had a negative test at the first or second survey after a positive test at the first survey). The highest rate of recovery (76.5%; 95%CI 50.1-93.2%) was observed in the age group of symptomatic 71-80 year old (**Table S2**). SARS-CoV-2 positivity overall (i.e. first and second survey combined) and at the first survey was more frequently associated with those aged 71-80 years (compared to 21-30 year olds, p-value = 0.01) (**Figure S4**). Being male was associated with COVID-19 positivity in the second survey (p-value = 0.04).

Analyses of the association between common comorbidities such as diabetes, hypertension, vascular diseases, respiratory diseases in asymptomatic and symptomatic individuals and the use of treatment for a number of different conditions with symptomatic infection showed no significant association (**Table S4** and **Table S5**).

### The role of asymptomatic individuals for transmission

The presence of a significant number of asymptomatic SARS-CoV-2 infections raises questions about their ability to transmit the virus. To address this issue, we conducted an extensive contact tracing analysis of the 8 new infections identified in the second survey (**Table 3**). Three of the new infections reported the presence of mild symptoms and did not require hospitalization. For Subject 1 we could not identify the source of infection. Subject 2 had contacts with four infected relatives who did not have any symptoms at the time of contact. Subject 3 reported contacts with two infected symptomatic individuals before the lockdown. Five of the 8 new infections showed no symptoms; Subjects 4 and 6 shared the same flat with symptomatic infected relatives. Subject 5 reported meeting an asymptomatic infected individual before the lockdown; Subject 7 did not report any contact with positive individuals and Subject 8 shared the same flat with two asymptomatic relatives. Notably, all asymptomatic individuals never developed symptoms, in the interval between the first and the second survey, and high proportion of them cleared the infection. The analysis of viral genome equivalents inferred from Ct (cycle threshold) data from real-time reverse-transcription PCR (RT-PCR) assays indicated that asymptomatic and symptomatic individuals did not differ when compared for viral PCR template recovered in the nasopharyngeal swabs (**Figure S5**). These results suggest that asymptomatic infections may play a key role in the transmission of SARS-CoV-2. We also found evidence that transmission can occur before the onset of symptoms, as detailed hereafter for a family cluster. Subject A (**Table S6**) was the first confirmed SARS-CoV-2 infection in the family, detected on February 22: the subject showed mild symptoms of the disease on February 22, was admitted to the Infectious Diseases unit on February 25 and subsequently discharged on February 29, with quarantine restrictions. The partner (Subject B) and children (Subjects C and D) tested positive on February 23 but showed only mild symptoms and did not require hospitalization. Subject A reported attending a family gathering three or four days before symptoms onset, together with a parent (Subject E) and three other siblings (Subjects F, G, and H). At that time, all of them were healthy. Nasal and throat swabs confirmed the presence of viral RNA in all family contacts. The transmission dynamics within this family clearly show that viral shedding of SARS-CoV-2 occurred in the early stages of infection and in the absence of symptoms. Furthermore, detailed analysis of contact tracing indicated that the relative risk of contracting the infection having an infected relative living in the same household gives an odd ratio of 84.5 (95% CI 16.8-425.4) (**Table S7**).

**Table 3.**
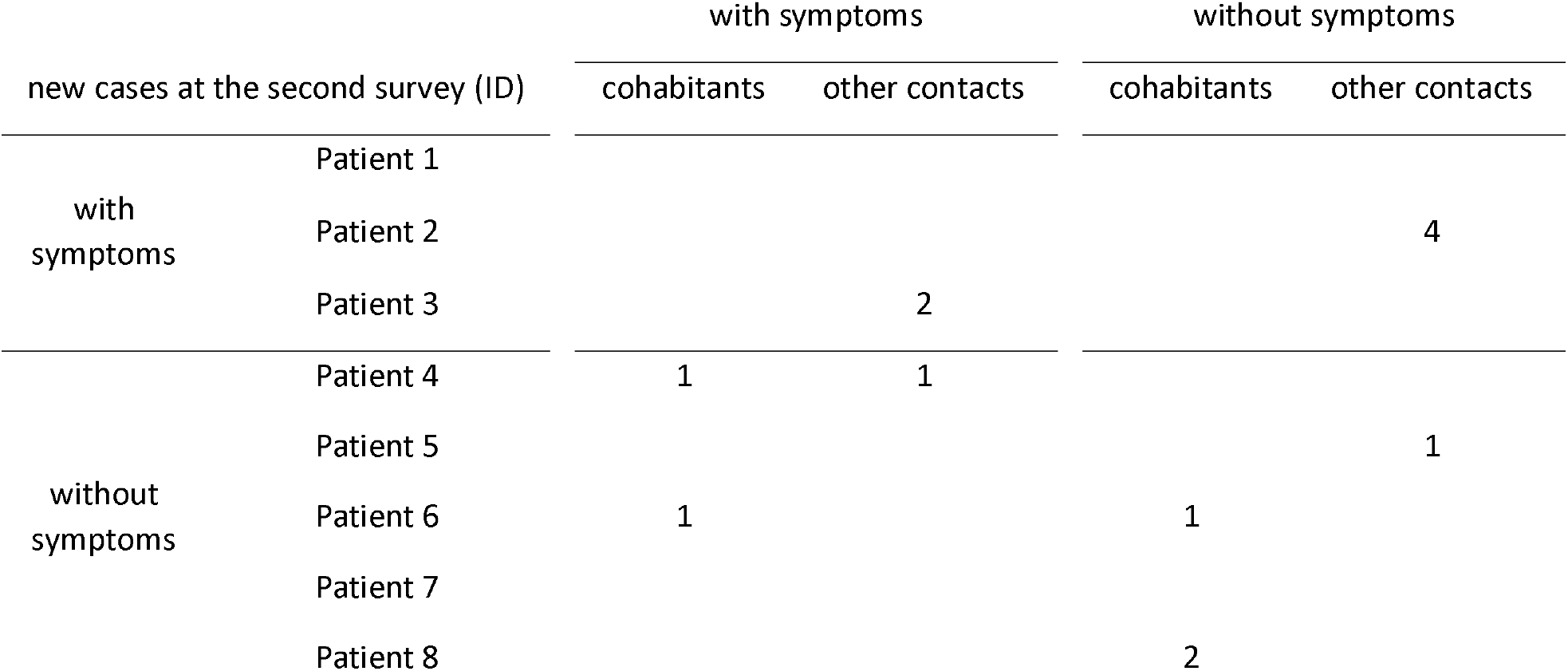
Contact tracing of all new infections detected in the second survey

| new cases at the second survey (ID) |  | with symptoms |  | without symptoms |  |
| --- | --- | --- | --- | --- | --- |
|  |  | cohabitants | other contacts | cohabitants | other contacts |
| with symptoms | Patient 1 |  |  |  |  |
|  | Patient 2 |  |  |  | 4 |
|  | Patient 3 |  | 2 |  |  |
| without symptoms | Patient 4 | 1 | 1 |  |  |
|  | Patient 5 |  |  |  | 1 |
|  | Patient 6 | 1 |  | 1 |  |
|  | Patient 7 |  |  |  |  |
|  | Patient 8 |  |  | 2 |  |

### Reconstructing transmission chains

We found that transmission was substantially reduced following the lockdown (**Figure S6**). From the inferred transmission pairs, we estimated a serial interval distribution with mean 6.90 days (95% CI 2.56-13.39) before lockdown (**Figure S7**) and 10.12 days (95% CI 1.67-25.90) after the lockdown. From the reconstructed chains of transmission, due to large initial clusters of transmission (**Figure S6**), we estimate an initial weekly effective reproduction number of 3.0 (95% CI 2.5-3.5) which declined to 0.14 (95% CI 0.0-0.29) by the end of the lockdown.

### Modelling point prevalence data

We used the prevalence estimates obtained in Vo’ at the first and second survey to calibrate a modified SEIR compartmental model of SARS-CoV-2 transmission that incorporates symptomatic and asymptomatic infections, virus detectability (in swabs) beyond the infectious period and the lockdown (**Figure S9**). We found that the model can reproduce the point prevalence data observed for symptomatic and asymptomatic infections (**Figure 3**). The mean and 95% credible intervals (CrIs) of the parameter estimates are given in **Table S8**. We estimated that on average 41-42% of the infections are asymptomatic and that the lockdown reduced SARS-CoV-2 transmissibility, on average by 89-99%, depending on the assumed initial value of ff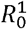 and on the duration of virus detectability (**Table S8**). The model suggests that at least 4.4% (95% CrI 3.6-5.3%) of the population of Vo’ was exposed to SARS-CoV-2 infection. The model suggests that values of the average duration of virus detectability beyond the infectious period in the range of 2-6 days (i.e. a total average duration of virus detectability including the infectious period of 4-8 days) better capture the central point prevalence estimates (Table S8, **Figure 3**). The estimated number of seeds suggest that SARS-CoV-2 was first introduced in Vo’ 1-3 generations of infections (depending on the assumed value of 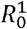) before, i.e. from the second half to the end of January 2020.

**Figure 3:**
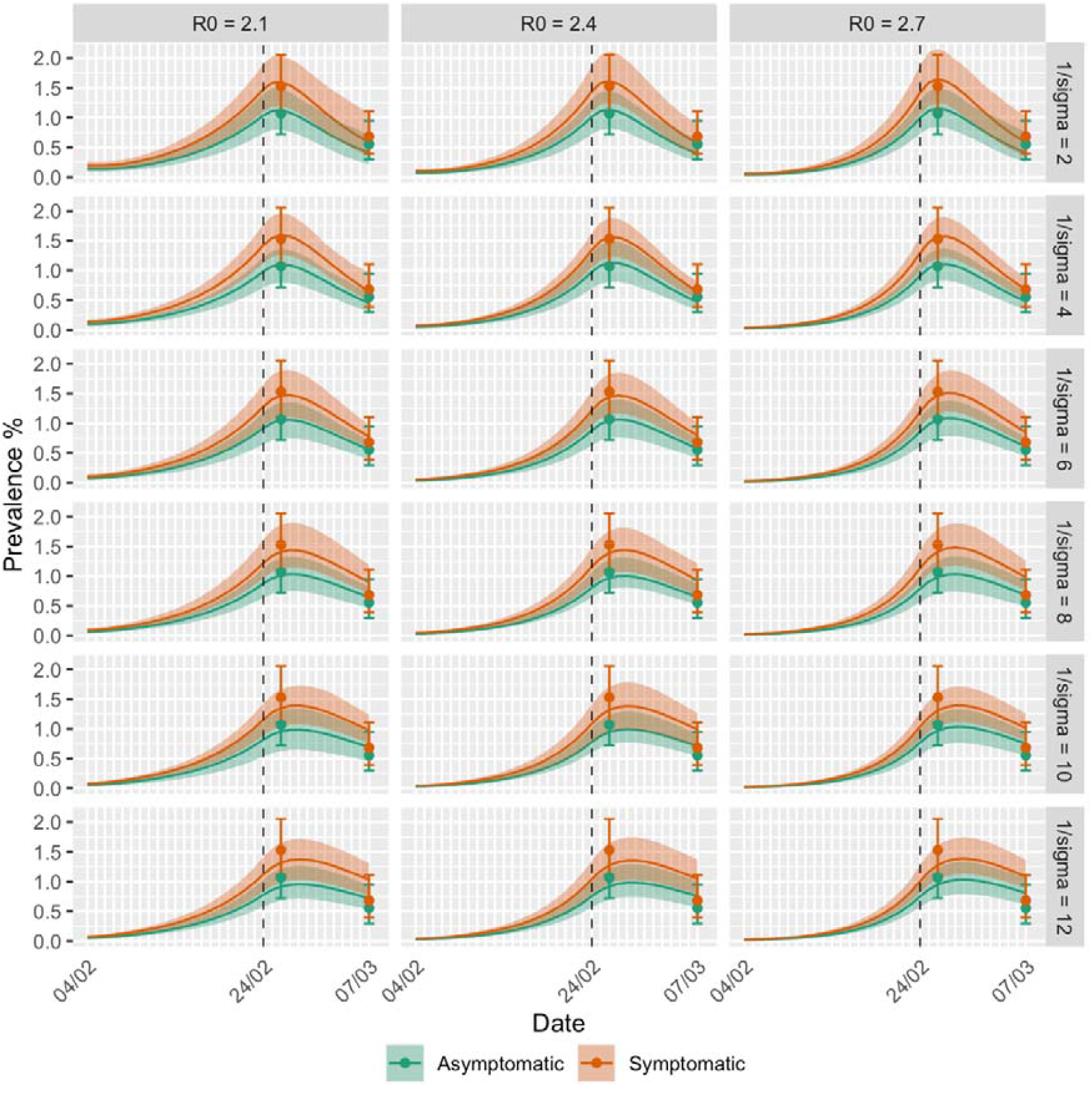
SARS-CoV-2 dynamics in Vo’ inferred from the observed prevalence data for symptomatic and asymptomatic infections in the first and second survey. Each sub-panel represents the model fit using the specified values of (the reproduction number before the lockdown) and 1/*σ* (the average duration of positivity beyond the duration of the infectious period). The dashed vertical line represents the time of quarantine implementation. The solid lines represent the mean and the shading represent the 95% CrI obtained from 100 samples from the posterior distribution of the parameters.

## Discussion

The result of the two surveys carried out in Vo’ provide important insights into the transmission dynamics of the virus, the contributions of symptomatic and asymptomatic infections to onward transmission, demonstrating that combining early active case detection and social distancing the transmission of SARS-CoV-2 was effectively suppressed in Vo’. The initial swab sampling was completed a few days after the first SARS-CoV-2 death was diagnosed, which was the earliest account of Italy’s epidemics. We found that 43.2% (95% CI 32.2-54.7%) of all confirmed SARS-CoV-2 infections across the two surveys were asymptomatic. Among confirmed SARS-CoV-2 infections, we did not observe significant differences in the frequency of asymptomatic infection in the different age groups (**Figure S10**). Recent studies found that while children are equally susceptible to SARS-CoV-2 infection as adults, their clinical progression is generally milder^13,14,15^. We found that none of the children who took part in the study tested positive for SARS-CoV-2 infection at either survey had a positive swab, despite at least 13 of them living together with infected family members. This is particularly intriguing in the light of the very high observed odd ratio for adults to become infected when living together with SARS-CoV-2 positive family members. However, this result does not mean that children cannot be infected by SARS-CoV-2. Nasopharyngeal swabs test for the presence of SARS-CoV-2 and can hence only detect active infection, not exposure. A cross-sectional serological survey would clarify the actual exposure of the whole population, including children’s exposure, to SARS-CoV-2. The pathogenesis of SARS-CoV-12 in young children is not well understood and children may have specific immune-regulatory mechanisms that contribute to milder disease or, alternatively, vaccinations or infection with other coronaviruses commonly transmitting in the youngest age groups may confer some level of heterologous protection against SARS-CoV-2^16,14^.

The analysis of the contacts of the 8 new infections identified in the second survey (**Table 3**), where we found that one had a history of close contact with asymptomatic individuals and two lived in the same household with asymptomatic relatives, suggests that asymptomatic infections can transmit the virus. The observation that the viral load in asymptomatic infections does not significantly differ from that of symptomatic infections further corroborates this hypothesis and the high frequency of asymptomatic infection detected across the surveys poses clear challenges for the control of COVID-19 in the absence of strict social distancing measures.

The serial interval estimates (mean 6.9 days; 95% CI 2.6-13.4) obtained from the individual-level data collected in Vo’ before the lockdown are in good agreement with the serial interval estimated in Lombardy, Italy and elsewhere^17,18^. During the lockdown we estimate a longer serial interval of mean 10.12 days (95% CI 1.67-25.90). Analysis of the transmission chains showed that clusters of infection occurred in the early phases of the epidemic and produced initial estimates of the effective reproduction number of mean 3.0 (95% CI 2.5-3.5). Awareness of COVID-19 transmission soon after the announcement of the first death and the town lockdown rapidly decreased the effective reproduction number below the threshold of 1 in the following weeks. These results are consistent with the initial reproduction number estimates (between 2.1 and 2.7, corresponding to a doubling time of 3-4 days) and the 89-99% drop in the reproduction number during the lockdown estimated from the fit of the dynamical model to the point-prevalence data observed in Vo’ at the first and second surveys. Notably, our results suggest that the dynamics as well as the duration of virus detectability in the swabs are comparable among symptomatic and asymptomatic infections, which is consistent with the viral load observed in symptomatic and asymptomatic infections in Vo’ and elsewhere^17,19^. Our results suggest that average durations of virus detectability in swabs between 4 and 8 days match more closely the observed prevalence of infection than longer durations (**Figure 3, Table S8**). This estimate is consistent with the observed distribution of the time from symptoms onset to confirmation which has mean 5.7 days (95% CI 1.6 – 12.3) (**Figure S6**). Our analysis suggests that at least 4.4% (95% CrI 3.6-5.3%) of the population of Vo’ was exposed to SARS-CoV-2. The interventions implemented in Vo’ substantially reduced the transmission of SARS-CoV-2 with unprecedented efficacy and demonstrate that COVID-19 suppression in similar epidemiological and demographic settings can be achieved. Enhanced surveillance and the early detection of SARS-CoV-2 transmission in places that have not yet been affected by the virus are key to control its spread and reduce the substantial public health, economic and societal burden posed by COVID-19 worldwide.

## Methods

### Study setting

The municipality of Vo’, in the province of Padua, Veneto region, Italy, is located about 50 kilometers west of Venice (**Figure 1A**). According to the latest land registry, Vo’ has a population of 3,275 individuals over an area of 20.4 square kilometers. Upon the detection of SARS-CoV-2 in a deceased resident of Vo’, we conducted an epidemiological study to investigate the prevalence of SARS-CoV-2 infection in the population as well as to assess the viral load of infected individuals. Sampling was conducted on the majority of the Vo’ population at two time points, the first during the days immediately after the detection of the first cases (21 – 29 February 2020) and the second one at the end of the two-weeks lockdown (07 March 2020) (Error! Reference source not found.B). For each resident we collected information on the sampling dates, the results of SARS-CoV-2 testing, demographics (e.g. age and sex), residence, health record (including symptom progression and hospitalization data, previous conditions and therapy taken for other illnesses), household size and composition, kinship ties and contact network. This information is available in the dataset spreadsheet (supplementary materials).

### Laboratory Methods

Nasopharyngeal swabs were collected by using flocked swabs in liquid-based collection and transport systems (eSwab^®^, Copan Italia Spa, Brescia, Italy). Total nucleic acids were purified from 200 μL of nasopharyngeal swab samples and eluted in a final volume of 100 μL by using a MagNA Pure 96 System (Roche Applied Sciences, Basel, Switzerland). Detection of SARS-CoV-2 RNA was performed by an in house real-time RT-PCR method, which was developed according the protocol and the primers and probes designed by Corman et al. targeting the envelope (E) (E_Sarbeco_F, E_Sarbeco_R, E_Sarbeco_P1) and RNA-dependent RNA-polymerase (RdRp: RdRp_SARSr-F, RdRp_SARSr-R, RdRP_SARSr-P1, and RdRp_SARSr-P2) genes of SARS-CoV-2. Real-time RT-PCR assays were performed in a final volume of 25⍰μL, containing 5⍰μL of purified nucleic acids, using One Step Real Time kit (Thermo Fisher Scientific, Waltham, MA, USA) and run on ABI 7900HT Fast Sequence Detection Systems (Thermo Fisher Scientific). The sensitivity of the E gene and RdRp gene assays was 5.0 and 50 genome equivalent copies per reaction at 95% detection probability, respectively. Both assays had no cross-reactivity with the endemic human coronaviruses HCoV⍰229E, ⍰NL63, ⍰OC43 and ⍰HKU1 and with MERS-CoV. All nasopharyngeal swab samples were screened with the E gene assay, followed by confirmatory testing with the RdRp gene assay. All tests were performed at the Clinical Microbiology and Virology Unit of Padova University Hospital, which is the Regional Reference Laboratory for emerging viral infections. After an initial period of dual testing by the National Reference Laboratory at the Italian Institute of Health (Istituto Superiore di Sanità), which demonstrated 100% agreement of results, the Regional Reference Laboratory received accreditation as Reference Laboratory for COVID-19 testing.

### Assessment of genome equivalents

Ct (cycle threshold) data from real time RT-PCR assays were collected for E and RdRp genes. Genome equivalent copies per ml were inferred according to linear regression performed on calibration standard curves. The interpolated Ct values were further multiplied by 100, according to the final dilution factor (1:100). Linear regression was calculated in Python3.7.3 using modules scipy 1.4.1, numpy 1.18.1, and matplotlib 3.2.1^21^. Genome equivalents distributions from the two genes, for both positive asymptomatic (35 individuals) and symptomatic (45 individuals), were compared with the Exact Wilcoxon-Mann-Whitney test. H_1_ hypothesis that the two distributions (asymptomatic vs. symptomatic) are different is rejected with p-values 0.6 and 0.2 for gene E and gene RdRp respectively.

### Reconstructing transmission chains

We used data on close contacts traced within the community and on household contacts derived from household composition data (available for all study participants) to impute chains of transmission and transmission clusters using the R package epicontacts^22,23^. This analysis included 166 cases and identified 120 directions of transmission. We inferred the date of symptoms onset for the subjects testing positive in the first survey but with missing onset date from a gamma distribution that was fitted to the observed time-lags from symptoms onset to sample collection available on those who tested positive at the first survey (Figure S6). Because access to immediate testing for those who developed symptoms was available from the start of the lockdown, we assumed that the distribution of the time-lag from the onset of symptoms to sampling in those confirmed in the second survey followed a normal distribution with mean and standard deviation equal to 1 day. Similarly, we used this latter distribution to infer the onset date for the subjects with confirmed infection in the second survey and missing onset date. We used the observed and inferred dates of symptoms onset alongside contact information to infer transmission chains within the sampled population. In turn, reconstructed transmission pairs were used to characterise the serial interval (the time between the onset of symptoms of the infector and the onset of symptoms of the infectee) and the effective reproduction number (the average number of secondary infections generated by the identified infectors). Weekly effective reproduction number estimates were calculated as the average number of secondary infections generated (over the whole study period) by subjects with symptoms onset during the same week, having stochastically assigned positive individuals with unidentified infectors to positive subjects with symptoms onset compatible with samples from the serial interval distribution and an infectious period of 2 days. The mean and 95% CI were calculated on 10,000 samples of the serial interval distribution.

### Mathematical modeling

The first survey occurred between 21^st^ and 29^th^ February 2020 and the second survey occurred on 7^th^ March 2020. In the model we assumed that prevalence was taken on the weighted average of the first sample collection, i.e. on 26^th^ February 2020 and on 7^th^ March 2020. We assumed that the population of Vo’ was fully susceptible to SARS-CoV-2 (S compartment) at the start of the epidemic. Upon infection, infected subjects incubate the virus (E compartment) for an average of 1/*δ* days before becoming infectious and able to transmit (I compartments). We assume that a proportion *p* of the infected population is asymptomatic (I_A_ compartment) and that the remaining proportion 1-*p* develops symptoms (I_S_ compartment). We assume a fixed infectious period of mean 1/*γ* days and that only infectious individuals (i.e. those in I_S_ and I_A_) contribute to onward transmission of SARS-CoV-2 but that the virus can be detected beyond the duration of the infectious period (this assumption is compatible with the hypothesis that transmission occurs for viral loads above a certain threshold but the diagnostic test can detect the presence of virus below the threshold for transmission). Compartments TP_S_ and TP_A_ respectively represent symptomatic and asymptomatic subjects who are no longer infectious but with a detectable viral load, and hence testing positive. Eventually, the viral load of all infections decreases below detection and subjects move into a test negative (TN compartment). We assume that symptomatic and asymptomatic infections are equally infectious (i.e. they contribute equally to the force of infection). The flow diagram of the model is given in **Figure S9**. We assume a step change in the reproduction number on the day that lockdown started. Before the implementation of quarantine the reproduction number is given by 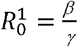 and we assume that it drops to 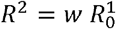 after the lockdown, where 1-w represents the percent reduction in *R*_0_ due to the intervention. We let *T*_*i*_ denote the number of subjects swabbed on survey *i* (*i* = 1,2) and let *P*_*Ai*_ and *P*_*Si*_ respectively denote the number of swabs testing positive among asymptomatic and symptomatic subjects, respectively. We assume that the number of positive swabs among symptomatic and asymptomatic infections on survey i follows a binomial distribution with parameters *T*_*i*_ and *π*_*Xi*_, where *π*_*Xi*_ represents the probability of testing positive on survey i for class X (= A,S), given by 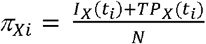 assuming perfect diagnostic sensitivity and specificity. The likelihood of the model is given by the product of the binomial distributions for symptomatic and asymptomatic subjects at times *t*_*i*_, *i* = 1,2. Inference was conducted in a Bayesian framework, using the Metropolis-Hastings Markov Chain Monte Carlo (MCMC) method with uniform prior distributions^24,25^. We fixed the average incubation period (1/δ) to 5 days and the average infectious period (1/γ) to 2 days, which gives an average generation time of 7 days^18,26^. We explored the following values of 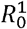: 2.1, 2.4, 2.7, which are compatible with a doubling time of 3-4 days, as observed in the Veneto region of Italy. We assumed that seeding of the infection occurred on 4 February 2020, i.e. 5 days before the date of symptoms onset of the first recorded case in Vo’. We explored different scenarios on the average duration of viral detectability beyond the infectious period and fixed 1/*σ* to be 2, 4, 6, 8, 10 and 12 days. We estimate the number of infections introduced in the population from elsewhere at time *t*_*0*_ (4 February 2020), p and 1 − *w*.

### Association between infection and age, sex and symptoms

We applied logistic regression to test the association between SARS-CoV-2 positivity (overall and at the first and second survey separately) with the age-group (10 years age bands, from 0 to 81+), sex (male, female) and the occurrence of symptoms (defined as the presence of fever and/or cough).

### Association with comorbidity and pharmacological therapies

We used Fisher’s exact test for comparing two binomial proportions to assess whether there is an association between the presence of symptoms for 39 confirmed COVID-19 cases resident in Vo’ and different types of comorbidities and treatments used. The analyses were repeated on the subset of patients who became negative at the second timepoint (results not shown). In an attempt to increase the power of the data, we increased the sample size by including additional 11 confirmed COVID-19 cases resident in other villages close to Vo’. None of these scenarios provided significant associations at the 5% level.

## Data Availability

The dataset is available as a supplementary file.

## Data availability

The dataset is available as a supplementary file.

## Code availability

The code developed to fit the mathematical model to the prevalence data is available at https://github.com/ConniCia/SEIR_Covid_Vo. The code developed for the reconstruction of the transmission chains and for the statistical analyses is available upon request.

## Ethical approval statement

The first sampling of the Vo’ population was conducted within the surveillance program established by the Veneto Region and did not require ethical approval; the second sampling was approved by the Ethics Committee for Clinical Research of the province of Padova.

## Materials & Correspondence

Please address correspondence to Prof Andrea Crisanti or Dr Ilaria Dorigatti.

## Author contributions

A.C. conceived the project with input from E.L., I.D.

I.D. conceived the modelling with input from N.M.F and C.A.D

E.L. coordinated data collection, curation and analyses.

E.F. coordinated the diagnostic team and facilities.

E.F., L.B., C.D.V., L.R., R.M., A.L., D.A.A., M.S., E.D., M.C.V., F.S., F.O., G.M., and M.T. performed laboratory testing on swabs and validated the results.

E.L., S.T., V.B., A.S., N.N., and S.C. analyzed the data, contributed to the discussion and commented on the manuscript.

A.R.B., I.D., C.A.D. performed statistical analyses.

C.C, L.C., N.M.F. and I.D. developed the mathematical model.

G.C.D., K.M.G., C.A.D and I.D. performed cluster analysis.

S.M., R.S., G.C., D.D., and L.F. organized sampling logistics, S.M. and R.S. performed swab samplings.

Imperial College London COVID-19 Response Team contributed to the discussion and background understanding of COVID-19 epidemiology.

A.C. and I.D. wrote the manuscript, with contribution from E.L., L.B., V.B. and C.A.D.

## Acknowledgements

This work was supported by the Veneto Region and was jointly funded by the UK Medical Research Council (MRC) and the UK Department for International Development (DFID) under the MRC/DFID Concordat agreement and is also part of the EDCTP2 programme supported by the European Union. I.D. acknowledges research funding from a Sir Henry Dale Fellowship funded by the Royal Society and Wellcome Trust [grant 213494/Z/18/Z]. L.O. and G.C.D. acknowledge research funding from The Royal Society. We thank F. Caldart, M.D., G. Castelli, M.D., M. Drigo, M.D., L. Fava, M.D., B. Labella, M.D., M. Nicoletti, M.D., E. Nieddu, M.D. for assistance in data collection and consistency check, F. Bosa and G. Rupolo from the Italian Red Cross for the support in patient samplings.

## Imperial College London COVID-19 Response Team

Kylie E. C. Ainslie, Marc Baguelin, Samir Bhatt, Adhiratha Boonyasiri, Olivia Boyd, Lorenzo Cattarino, Constanze Ciavarella, Zulma Cucunubá, Gina Cuomo-Dannenburg, Bimandra A. Djafaara, Christl A. Donnelly, Ilaria Dorigatti, Sabine L. van Elsland, Rich FitzJohn, Seth Flaxman, Han Fu, Katy A.M. Gaythorpe, Will Green, Timothy Hallett, Arran Hamlet, Katharina Hauck, David Haw, Natsuko Imai, Ben Jeffrey, David Jorgensen, Edward Knock, Daniel Laydon, Thomas Mellan, Swapnil Mishra, Gemma Nedjati-Gilani, Pierre Nouvellet, Lucy C. Okell, Daniela Olivera, Kris V Parag, Steven Riley, Hayley A. Thompson, H. Juliette T. Unwin, Robert Verity, Michaela Vollmer, Patrick G.T. Walker, Caroline E. Walters, Haowei Wang, Yuanrong Wang, Oliver J Watson, Charles Whittaker, Lilith Whittles, Xiaoyue Xi, Neil M. Ferguson.

